# My Early Relational Trust-Informed Learning (MERTIL) for Parents: A study protocol for a brief, universal, online, preventative parenting program to enhance relational health

**DOI:** 10.1101/2022.07.14.22277633

**Authors:** Jessica Opie, Leesa Hooker, Tanudja Gibson, Jennifer McIntosh

## Abstract

**Background:** Early relational health is a key determinant of childhood development, while relational trauma in the parent-infant dyad can instigate a cascading pattern of infant risk. Fortunately, early relational trauma is detectable and modifiable. In 2018, Australian Maternal and Child Health (MCH) nurses participated in MERTIL (My Early Relational <u>Trauma</u>-Informed Learning), a program to identify and prevent relational trauma. Program evaluations revealed nurses felt competent and confident to identify and respond to relational trauma; however, response capacity was inhibited by inadequate parent referral options. In response, MERTIL *for Parents* (My Early Relational <u>Trust</u>-Informed Learning) was developed, which is an online, evidence-based, self-paced parenting program that focuses on enhancing parental knowledge of relational trust and its significance for infant development. This low-cost, accessible prevention resource targets emerging relational concerns to reduce later service system engagement. The potential for universal preventative online programs that target parental and relational wellbeing remains under-explored. This paper reports on a protocol for implementing a MERTIL *for Parents* pilot study describing nurses’ and parents’ perspectives on program feasibility and efficacy.

**Methods:** This study is a mixed-methods, parallel-armed, uncontrolled, repeated measures design. We aim to recruit 48 Australian MCH nurses from the states of Victoria and New South Wales. These nurses will in turn recruit 480 parents with a child aged 0-5 years. All parents will receive MERTIL *for Parents*, which entails a 40-minute video, tipsheets, worksheets, and support resources. Parent data will be obtained at three periods: pre-program, program exit, and program follow-up. Nurse data will be collected at two periods: parent recruitment completion and program follow-up. Data collection will occur through surveys and focus groups. Primary parent outcomes will be socioemotional assessments of program efficacy. Nurses and parents will each report on study program feasibility.

**Discussion:** This protocol describes the feasibility and efficacy of a new online parenting program, MERTIL *for Parents*, with pilot field studies commencing in August 2022. We anticipate that this resource will be a valuable addition to various child and family services, for use in individual support and group work.

## Study Protocol Introduction

Early relational health is a key determinant of childhood development, while relational trauma in the parent-infant dyad can instigate a cascading pattern of infant risk. Fortunately, early relational trauma is detectable and modifiable. In 2018, Australian Maternal and Child Health (MCH) nurses based in the state of Victoria participated in MERTIL (My Early Relational Trauma-Informed Learning), a program to identify and prevent relational trauma. Program evaluations revealed nurses felt competent and confident to identify and respond to relational trauma; however, response capacity was inhibited by inadequate parent referral options for those with sub-clinical presentations. In response, MERTIL *for Parents* (My Early Relational Trust-Informed Learning) was developed. MERTIL *for Parents* is an online, evidence-based, self-paced parenting program that focuses on enhancing parental knowledge of relational trust and its significance for infant development. This low cost, accessible prevention resource targets emerging relational concerns to reduce later service system engagement. The potential for universal preventative online programs that target parental and relational wellbeing, as well as the development of early relational trust, remains under-explored.

This protocol describes the feasibility and efficacy of a new online parenting program, MERTIL *for Parents*, describing MCH nurses’ and parents’ perspectives on program feasibility and efficacy, with pilot field studies soon to commence. This study is a mixed-methods, parallel-armed (i.e., MCH nurses and parents), uncontrolled, repeated measures design. We aim to recruit 48 MCH nurses who will in turn recruit 480 parents with a child aged 0-5 years (inclusive) displaying early signs of relational trauma. All participants will receive access to the MERTIL *for Parents* program, which entails a 40-minute video, tip sheets, worksheets, and support resources. Parent and nurse data collection will occur through surveys and focus groups. Primary parent outcomes will be socioemotional to examine program efficacy; such outcomes will include child-parent relationship quality and parental reflective functioning. Nurses and parents will each report on study program feasibility. It is anticipated that this resource will be a valuable addition to a wide range of child and family services, for use in individual support and group work.

## Background

Children develop in a relational context with everyday parent-child interactional patterns contributing to their *relational health* (1). Relational health in the infant-parent dyad is construed as parental security, trust, responsiveness, and sensitivity, which together form the cornerstone of socioemotional development. Alternatively, *relational trauma* presents ongoing and accruing disruptions to the infant’s context of early care. Unsurprisingly, relational health and relational trauma impact upon an array of child developmental domains, instigating a cascading pattern of child resiliency or risk. Importantly, relational health elements, including attachment organisation and parental sensitivity (2-4) are modifiable, for better or worse. With this knowledge, promoting positive caregiving via psychoeducation and increased parenting capacity is important, enabling parents to promote child development, thereby affording their child the best start in life. Early parenting programs have been developed as a result.

### Preventative Parenting Programs

Preventative parenting programs can be universal, selective, or indicated. Universal programs are those available to all parents; selective programs target parents displaying above average risk; and indicated programs target parents showing signs or symptoms of emerging disorder (5). Research has shown universal programs are well suited to mental health promotion, offering significant preventative public health benefit (6).

#### Universal face-to-face parenting programs

Universal parenting skills-based programs have historically been presented through varied in-person modes, including face-to-face lectures, workshops, and groups. Program examples include CANparent (7) and *ACT Raising Safe Kids Program* (8). Such programs have been shown to increase both child and parent mental health among an array of domains such as parental self-efficacy, mental wellbeing, satisfaction, and child internalising and externalising presentations (e.g., 7, 8, 9).

Despite the availability and evidence supporting these in-person parenting programs, many families, particularly those experiencing numerous vulnerabilities, face multiple barriers in accessing these programs (10, 11). For parents who live in regional centres and rural or remote locations, attending in-person programs, typically city-based, is an accessibility obstacle. Transportation access is also an issue for in-person parent service participation (12, 13). Such programs are additionally cost prohibitive to many due to their high in-person contact hours with professionals (14). The inflexible structure of such programs and rigid delivery approaches can be perceived as stigmatising by the individuals these programs are intended to reach. Capped attendance due to physical room size, occupational health and safety concerns, and the limited number of facilitators running sessions provide further shortcomings. Parent logistical barriers, such as managing various daily work and lifestyle demands have resulted in low in-person parent program enrolment and poor program attendance for enrolees (e.g., 5, 15-17). Compounding these limitations, the COVID-19 pandemic saw an abrupt stop to most in-person treatment programs due to lockdowns and quarantine, severing preventative program resources at a time of heightened psychological vulnerability for parents and children alike (18).

#### Universal online parenting programs

Online or app-based program delivery provides an avenue to address these limitations, provided intended users have adequate technology access, reliable internet connection, and are digitally literate. Online universal parenting programs can be either entirely or partially self-guided. In partially self-guided programs, clinician support is provided at varying frequencies via online, phone, or in-person modalities (19). Prior meta-analytic research on the efficacy of self-guided and partially self-guided online parenting programs has reported benefits at the parent, child, and dyadic level. Specifically, at the parent level, programs have been shown to enhance elements such as positive parenting perceptions, behaviours, satisfaction, efficacy, and confidence, while lowering parental mental health problems (e.g., anxiety, anger, depression, stress) and negative parenting practices (20-25). At the child level, programs decrease emotional and behavioural problems (21-23, 25), while at the relational level, they can reduce negative parent-child interactions (23), negative parental discipline strategies, and parental conflict (24).

While both self-guided and partially self-guided online parenting programs are efficacious, the formation and implementation of wholly self-guided, universal, digital parenting programs has recently gained momentum (e.g., 15, 26). The rapid uptake of such programs is particularly due to the ease of administration, scalability, sustainability, low-cost, and program standardisation. Further rationale for the uptake of online program delivery is due to the ubiquitous nature of internet access, allowing widescale dissemination. For example, 99% of Australians and 91% of Australian households have internet access (27), making this a delivery method that can reach most of the population, thus making these programs highly accessible. The self-directed nature of online programs may further allow parents to flexibly complete these programs in the security of their homes, thereby overcoming possible judgement and stigma felt from participating in in-person programs. Such benefits may in turn reduce costs and barriers to program engagement and enhance program reach.

Due to their significant preventative potential, online parent resources may reduce the need for later intervention. This may occur by supporting and motivating parents with new knowledge and behavioural skills to independently enhance their parent-child relationship, in turn reducing later burden on service systems. Public health initiatives such as these may empower parents to alter relational trajectories early on, within a preventative approach that is parent-led and strengths-based. This may assist to disrupt emerging disadvantage before reaching pathological thresholds. Equitable access through online platforms has become more important, particularly in the face of the COVID-19 global crisis. Further, evidence shows that these programs promote help-seeking (28). Finally, from a research perspective, fully online content allows for the collection and tracking of unique data on training progress, completion times, and premature program cessation. Such data can be linked to outcomes and inform program improvements.

### Knowledge Gaps

While numerous clear benefits exist in relation to online, self-delivered, universal parenting programs, gaps remain. Currently, there are few brief self-help, psychoeducational, program options (in-person or online) suitable at the broad community level for parents where enhanced awareness of their role in promoting early relational trust may assist with prevention of relational trauma, especially in the face of challenge. Second, most parenting resources in this domain exist in the context of in-depth indicated parenting interventions (e.g., Parent-Child Psychotherapy; 1). While these indicated programs are essential for dyads in need of targeted treatment, there is a clear need for preventative education resources for parents in the wider community wishing to enhance their understanding of the parent-child relationship. This could act as an initial independent healthcare measure and reduce the need for later targeted measures. Further, outcomes of online parenting programs tend of focus on parents and, to a lesser extent, child outcomes (21, 24). Few studies have reported dyadic outcomes and even fewer have reported familial outcomes associated with online parent program participation (e.g., 23). Finally, despite MCH nurses completing MERTIL and providing parenting support, they do not have a recommended suite of online parenting programs within their toolbox to assist clients (29). MCH service practice guidelines focus on health promotion, child development and to a lesser extend parent mental health; however, nurses need evidence-based interventions to enhance dyadic relationships. This study aims to address these resource and knowledge gaps.

## Methods

### Study aims

This paper reports on a protocol for the pilot implementation of a new self-directed, online parenting program (MERTIL *for Parents)* within the Maternal and Child Health (MCH) and allied Early Childhood sector. The aims of this study are twofold:

1. Determine the feasibility of MERTIL *for Parents:*
  i. For MCH nurses and allied health professionals: Examine the programs acceptability and utility for enhancing parent understanding of early relational health; assess the practical dimension of implementing the program within a busy service; and explore population reach.
  ii. For parents: Examine program acceptability, efficacy, and utility.
2. Evaluate the short-term efficacy of the MERTIL *for Parents* program for parents across selected parental caregiver knowledge, perceptions (e.g., parenting confidence & competence), and behavioural domains (e.g., professional help-seeking amenability).

### Intervention development

MERTIL *for Parents* is a new universally available, self-directed, online psychoeducation resource for Australian parents focused on promoting parental knowledge and understanding of relational trust and its significance for infant development. This program is informed by evidence from attachment theory, developmental neuroscience, and psychoanalysis. As a preventative early mental health resource, MERTIL *for Parents* aims to enhance infant-parent relational trust and relational awareness, in turn modifying parents’ attachment security-oriented beliefs, attitudes, and behaviours. MERTIL *for Parents* builds upon prior research by the authors. In 2018, over 1700 Australian MCH nurses from the state of Victoria participated in MERTIL (My Early Relational <u>Trauma</u>-Informed Learning), a 14-hour self-directed, interactive, professional development program for MCH nurses to identify and prevent relational trauma. The MERTIL program evaluation from the 2018 Victorian cohort revealed that MERTIL enhanced competence and confidence in nurses’ identification of relational trauma (30). However, capacity to respond to such identified early relational trauma was inhibited by inadequate referral options for parents, particularly in rural and remote settings. More broadly, it was identified that the potential for universal online programs that target the development of early relational trust remains under-explored (30). In response, MERTIL *for Parents* was developed, replacing the *trauma* in MERTIL for *trust*. See Fig 1 for the MERTIL *for Parents* logo.

**Fig 1.**
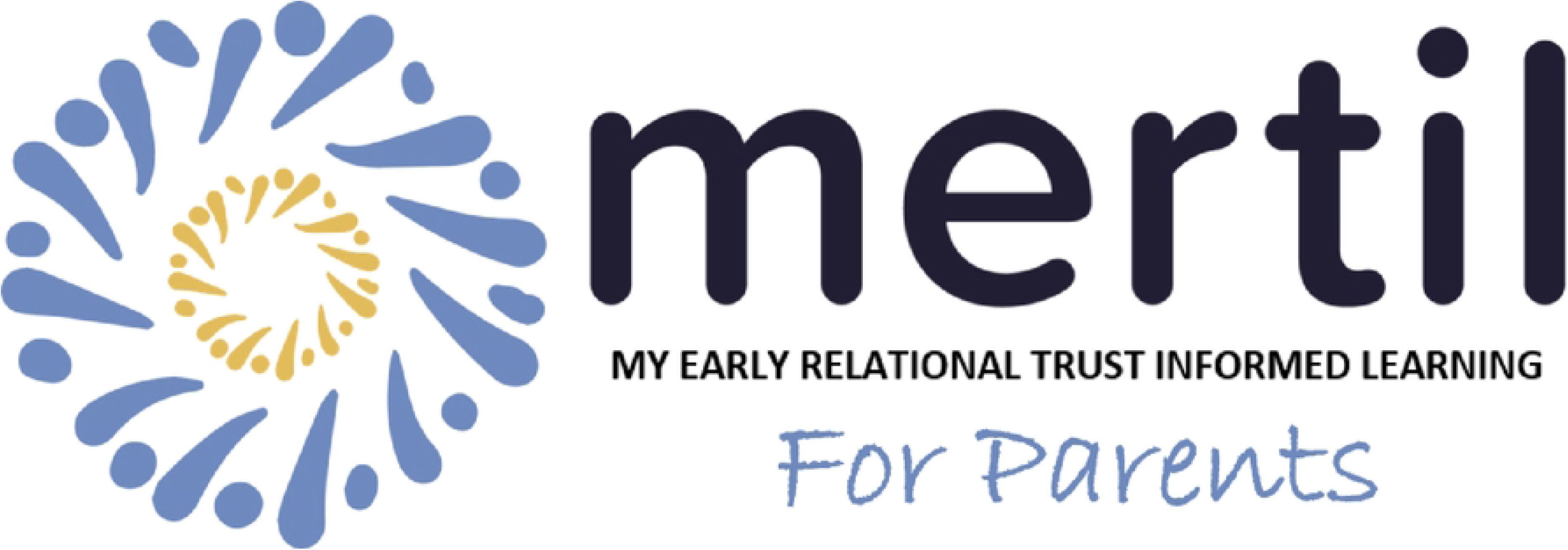
MERTIL *for Parents* logo.

### The Intervention

#### Intervention Content

MERTIL *for Parents* is accessed via the internet and can be viewed on any online platform (i.e., phone, computer, or tablet). The primary component of MERTIL *for Parents* comprises a 40-minute pre-recorded video. Video content consists of four ‘chapters’, with each chapter approximately nine-minutes in duration. See Table 1 for a description of the MERTIL *for Parents* video content. Secondary program components include downloadable parent tip sheets, parent worksheets, and professional support contacts. All secondary program elements will be developed by the study authors. See Table 2 for additional downloadable program resources. Participants progress through the varied program aspects at their own pace.

Table 1. MERTIL *for Parents* video content.

Table 2. Non-video MERTIL *for Parents* content

#### Program engagement

Numerous steps were taken to ensure MERTIL *for Parents* is highly engaging, in order to 1) enhance program enrolment, 2) deliver memorable content, and 3) maximise successful program completion. MERTIL *for Parents* was intentionally designed as a very-brief parenting program, to counter the high dropout rates that have been observed in other online parenting programs (31). The rationale for MERTIL *for Parents’* length was to introduce parents to the pivotal elements of relational trust, in a manageable, user-friendly, and time-efficient fashion that could be completed in one sitting or across several brief stints. A production team and animator translated the content into an engaging, animated format. See Fig 2 for screenshots of the MERTIL *for Parents* content.

**Fig 2.**
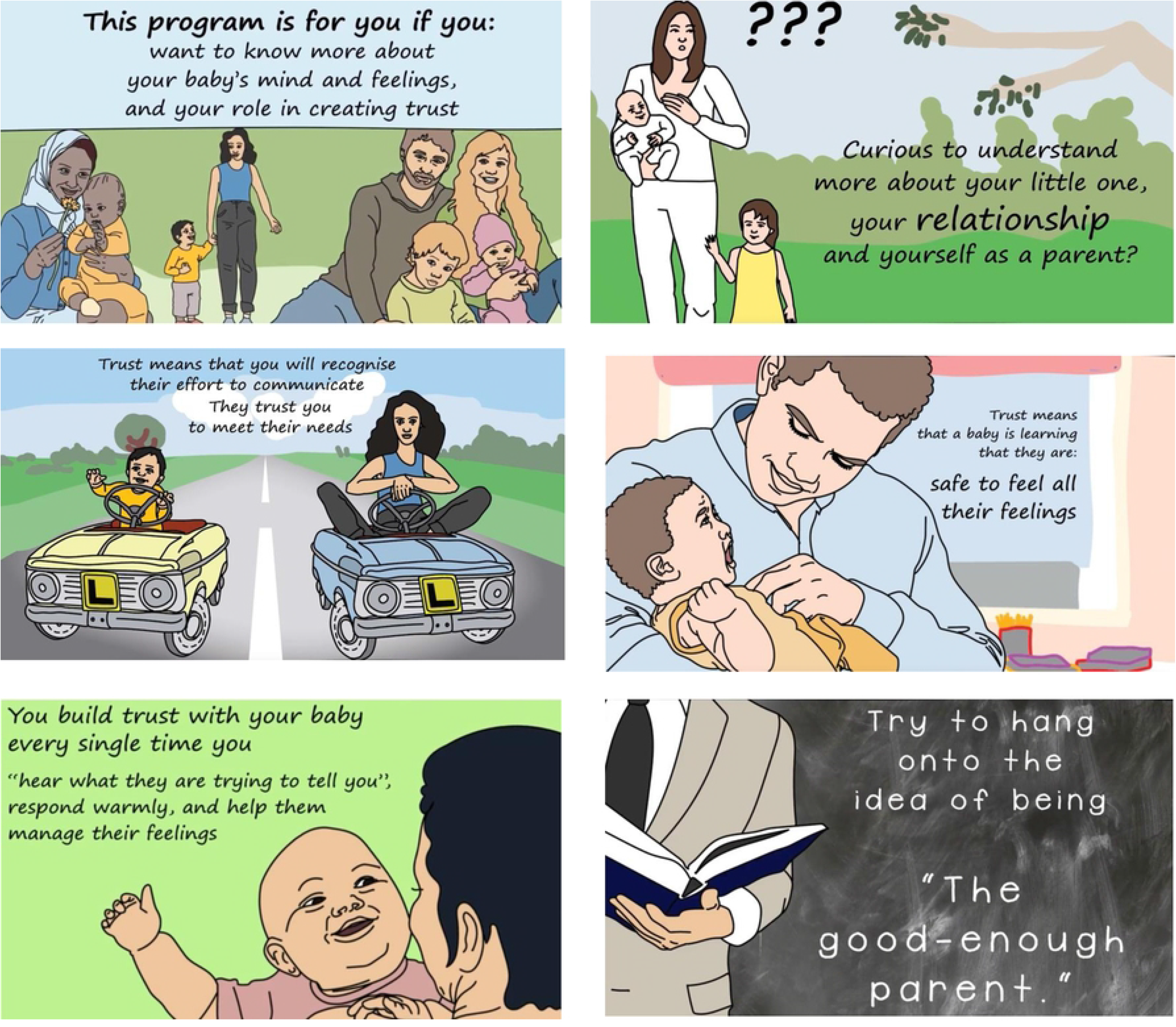
Screenshots from MERTIL *for Parents* video content.

##### Program content & delivery

To ensure the evidence-based narrative content was engaging and well received, content included:

1. Occasional moments of humour to balance the program’s occasional heavy content.
2. Lay language to ensure the program’s content was successfully communicated to a varied parent audience,
3. An emphasis on safety, relatability, and acceptance; to this end, the narrator’s voice is warm, calm, non-blaming, and reassuring.

##### *Co-designed nature of* MERTIL *for Parents*

The MERTIL *for Parents* website (www.mertil.com.au) and all content within, was co-designed and co-developed alongside consumers, child infant mental health clinicians, and MCH nurses alike, from program commencement to completion. This collaborative process enabled the voice of ‘would be’ end-user parents and professional referrers to be heard; prior research has identified this co-design process as enhancing engagement, efficacy, and research applicability (32, 33). Researchers and clinicians engaged in a multiphase feedback process, culminating in the resultant MERTIL *for Parents* program.

### Design

MERTIL *for Parents* will be examined via a parallel armed (i.e., nurses & parents), uncontrolled, mixed-methods, repeated measures study design. Nurses will complete two assessments (post-parent recruitment and three-month post-program) and parents will complete three assessments (pre-program, program exit, and three-month post-program). All nurse and parent data collection and completion will be conducted independently and in parallel.

### Participants

Eligible study participants will be twofold: 1) Australian MCH nurses, and 2) Australian parents. See Fig 3 for an outline of all study participants.

**Fig 3.**
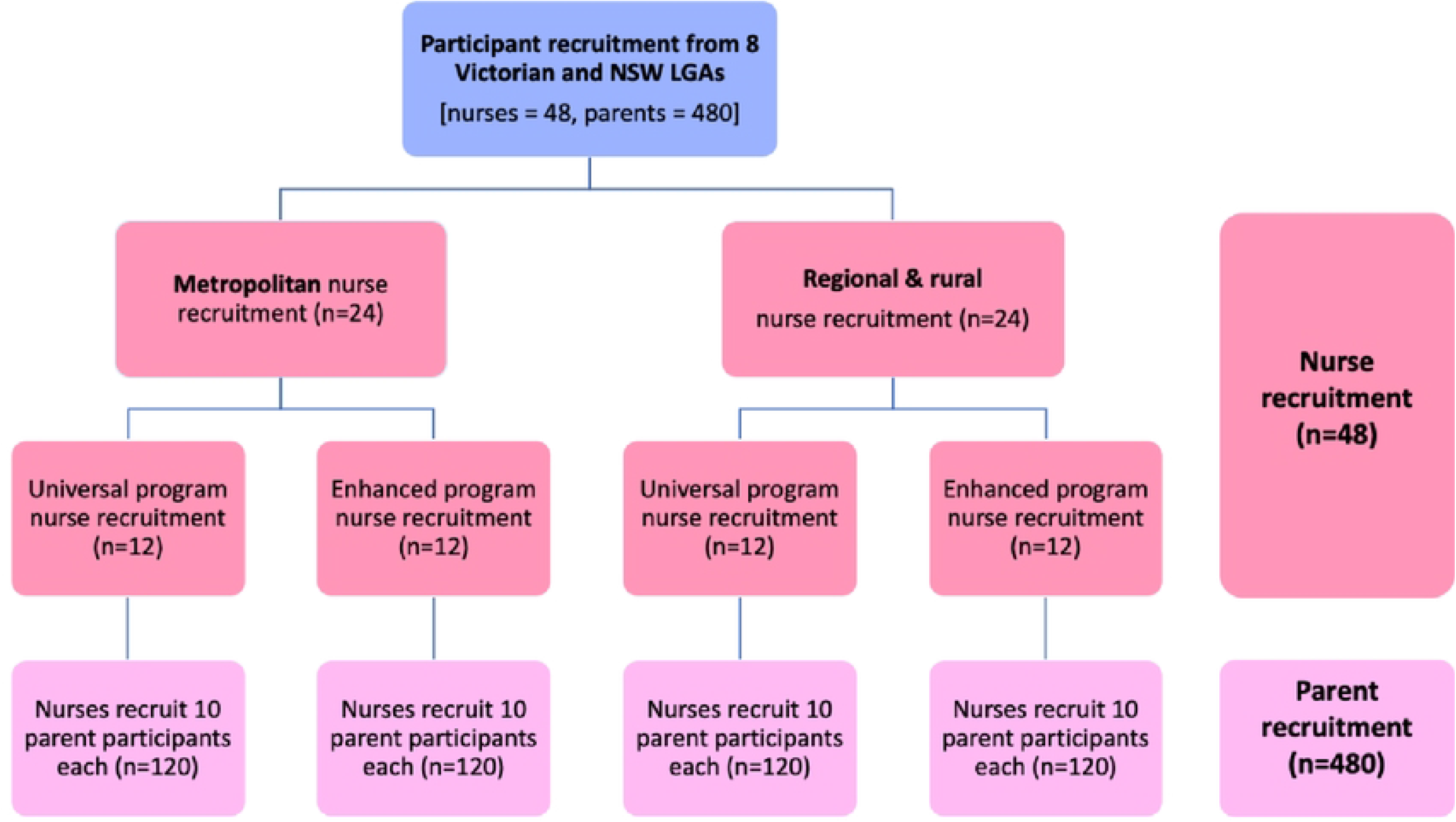
MERTIL *for Parents* recruitment diagram.

### Eligibility criteria

#### Nurses

Nurses will i) have completed MERTIL, and ii) work at a participating pilot MCH site, in metropolitan and regional/rural Victorian and New South Wales Local Government Areas (LGAs) and iii) be able to make a follow-up phone call to each of the parents they refer to the program, to gather feedback. Three programs sit within the MCH umbrella: 1) Universal service, 2) Enhanced service and 3) 24-hour MCH Telephone Line (29). Participants will be comprised of Universal and Enhanced services MCH nurses. The Universal MCH service is available to all families, while the Enhanced MCH service supports families who meet strict ‘at-risk’ criteria, with limited places. Study participation invitations will be sent to MCH nurses who work in both the Universal and Enhanced MCH services. For nurses who decide to participate, free access to MERTIL (My Early Trauma-Informed Learning) will be provided for the duration of the pilot study. This is the training program MCH nurses completed in 2018 and will allow them to refresh their knowledge of and skills in identifying early relational trauma, if necessary. Nurses will also have ongoing access to MERTIL *for Parents*.

#### Parents

Consenting parent participants will: i) be the parent of a child 0-5 years (inclusive) or currently pregnant, ii) be recommended to participate in the program by an MCH nurse from a participating pilot site, iii) aged 18 years and over, iv) currently reside in Australia, and v) feel comfortable to complete questionnaires and program materials in English.

### Sample size/power calculation

Power analysis was conducted using the G*Power 3.1 software to determine the required sample size for this study (34). A single group within study design would require 199 participants to identify a small to moderate statistically significant effect (.20) at *p*<.05, with 80% power. To account for parent participant attrition, we aim to recruit approximately 480 parents.

#### Sample

We aim to recruit 48 MCH nurses, consisting of 24 MCH nurses from the Universal MCH service and 24 MCH nurses from the Enhanced service. These nurses will in turn recruit the above-mentioned 480 parents.

### Recruitment of study populations

#### Nurse

The authors will recruit through their existing MERTIL MCH nurse networks, ensuring a representative sample of MCH nurses from differing Australian geographic locations. Participating MCH nurse sites will include four Victorian and New South Wales metropolitan LGAs and four Victorian and New South Wales regional/rural LGAs, based on pre-existing nurse networks. To further ensure representativeness, recruitment will include MCH nurses from the Universal and Enhanced MCH services. All participating MCH nurses will provide written online consent to their study participation.

#### Parents

Parents who meet inclusion criteria will be informed about the study by their MCH nurse during a visit to the Universal service (any of the scheduled visits from the initial post-birth home visit, consultations at 2, 4, & 8 weeks; 4, 8, 12, & 18 months; 2 & 3.5 years), or a visit from the Enhanced service. The MCH nurse will explain MERTIL *for Parents* and the associated research project, and if the parent verbally expresses interest, nurses will direct them to the MERTIL *for Parents* website to register for the study and to access the materials (www.mertil.com.au). Nurses will provide parents with a printed flyer with MERTIL *for Parents* details to aid this process. For parents who verbally agree to participate, MCH nurses will follow-up with a phone call or email, to gauge parents’ responses to the program.

During the study pilot phase, all individuals who arrive at the MERTIL *for Parents* website landing page will be invited to join the research. Specifically, parents will be met with a webpage popup with a brief research overview and the option to hear more about the study. If they select to hear more about the research, they will be automatically prompted to enter their first name and email address. This will trigger emailing of consent forms. Once the completed forms are submitted, the pre-program parent survey will be automatically emailed to the participant. Completing this survey will grant parents access to the MERTIL *for Parents* program content.

### Procedure

#### Nurses

Following informed consent, MCH nurses will attend a 120-minute web-based information and training session on MERTIL *for Parents*. This session will provide a study overview to promote program familiarisation, discuss parent program engagement, and explain the implementation options for parents accessing both the Universal and Enhanced MCH services. Immediately following this session, nurses will be emailed and asked to complete an online survey to report their first impressions of MERTIL *for Parents* regarding factors such as perceived program content, length, and suitability.

Nurses will be asked to make contact (phone call preferred) with parents they referred to the program, within two weeks of recommending MERTIL *for Parents*. On completion of all parent recommendations, a follow-up email survey will be conducted, assessing program use, perceived parent impacts, program engagement, ideas for program refinement, and wider implementation. Online focus group interviews will follow to elaborate on survey responses, assess nurses’ views and experiences of the program, potential barriers to implementation, and to identify further uses and applications of MERTIL *for Parents*. These elements will ensure MERTIL *for Parents’* co-designed nature endures. See Fig 4 for the MCH nurse data collection flow diagram.

**Fig 4.**
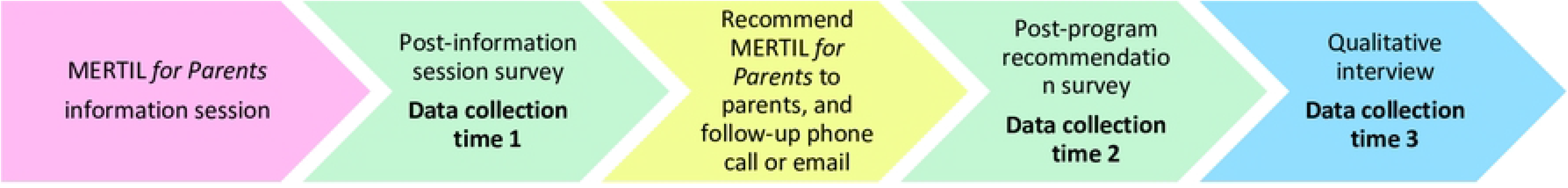
Flow diagram of MERTIL *for Parents* MCH nurse data collection timeline.

#### Parents

During the study pilot phase, all individuals who arrive at the MERTIL *for Parents* website landing page will be welcomed to the MERTIL *for Parents* pilot study and advised that the program is open to parents who are willing to participate in the research (but will be open to all following our test phase). They will be invited to read more about the study. If they consent, parents will provide their contact details to be emailed the Participant Information Statement and Participant Informed Consent Form. Once these forms are complete, the pre-program parent survey will be automatically electronically emailed to the parent participant.

The brief 10-minute survey before, after, and at 3-months follow-up comprises two sections: 1) close-ended questions, and 2) open-ended questions. Upon completion of Survey 1, participants will be able to access the MERTIL *for Parents* program. Participants have program access for 2 months from consent completion. Immediately upon MERTIL *for Parents* main video completion, Survey 2 will be sent. As needed, survey reminders will be sent via email at 7- and 14-days after the initial survey. If participants do not complete surveys after these communication bids, no further contact will be made. Three-months after completion of MERTIL *for Parents*, a follow-up survey will be sent to explore sustained change; consenting parents will participate in a qualitative interview, to further identify lived experience, program strengths and weaknesses. See Fig 5 for an outline of parent data collection points.

**Fig 5.**
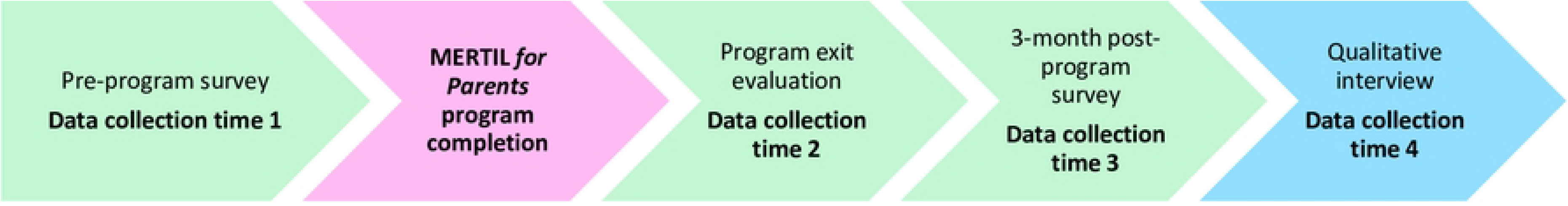
Flow diagram of MERTIL *for Parents* parent data collection timeline.

During this research and piloting phase, MERTIL *for Parents* will be free of charge to parents and caregivers who choose to participate in the research. Following this piloting phase and program refinement, MERTIL *for Parents* will be made broadly available.

### Measures

Our primary outcome will be to determine the efficacy of MERTIL *for Parents* in enhancing socioemotional development. Secondary outcomes include evaluating the feasibility of implementing MERTIL *for Parents* within the Victorian and New South Wales MCH services.

#### Parents

##### Baseline and three-month post-program questionnaires

To evaluate program efficacy, participants will complete identical self-report measures pre-program and three-month post-program (see Table 3); noting that demographic questions would have been completed pre-program and will not be repeated. On both occasions parents will receive surveys through a secure weblink sent to their email address. Program efficacy questions will be both quantitative and qualitative in nature. Program efficacy will not be assessed at program exit as not enough time would have elapsed for meaningful parent change. All included measurement items were derived where possible from validated scales, single items from validated scales, and/or items from established clinical cohort studies (i.e., Australian Temperament Project; Longitudinal Study of Australian Children). Table 3 displays outcomes and associated measures that will be used to evaluate MERTIL *for Parents* pre- and post-program.

Table 3. Pre- and post-MERTIL *for Parents* self-report outcomes, measures, & completion time.

1. Parental distress (depression & anxiety): The *Patient Health Questionnaire-4* (PHQ4; 35) will measure parent distress, with the two subscales measuring depression and anxiety.
2. Parenting practices: Parenting practices will be assessed by three parenting dimensions: hostile parenting (4 items), parental warmth (4 items), and parental self-efficacy (1 item; 36, 37).
3. Social support: the *Maternity Social Support Scale* (MSSS; 38) will determine parents’ perceptions of support. Item wording will be modified so items are applicable to all caregivers.
4. Intimate partner safety: The *Family Law DOORS* (FL-DOORS; 39) will measure partner safety via one item.
5. Parenting experiences: Select items from the *Perinatal Emotional Growth Index* (PEGI; 40) will assess parenting experiences. Seven items will assess parental confidence (2 items), parenting satisfaction (1 item), parental agitation (2 items), and general reflective capacity (2 items).
6. Relational attunement: The Interest-Curiosity subscale from the *Parental Reflecting Functioning Questionnaire* (PRFC; 41) will measure parents’ interest and curiosity in their child’s mental states. Two further study-generated questions will assess a parent’s capacity to respond to their child’s cues: 1) ‘I have built trust with my child’ and 2) ‘I know how to respond to my child’s signals’. Response ranges from 1 (completely true) - 5 (completely untrue).
7. Parent-child attachment: The Quality of Attachment subscale from the *Maternal Postnatal Attachment Scale* (MPAS; 42) will measure the quality of parent-child attachment. Item wording will be modified so items are applicable to all caregivers.
8. Professional help-seeking amenability: A study-specific item was developed by the authors (JM & JO) to assess parent’s help-seeking capacity: ‘I feel able to find and use support from other services in the community?’ with ratings from 1 (not at all) -10 (definitely).

Parents’ online program usage data will further be automatically collected via the MERTIL *for Parents* website. This will record program engagement and completion elements such as 1) program completion rates, 2) number of access occasions, 3) program components most/least accessed.

In addition to the above items, in the pre-program and three-months post-program questionnaires, parents will be asked open-ended free-text response questions relating to the program. Pre-program questions will cover hopes for learning, and three-month post-program feedback questions will include items on program components that parents found most useful.

Embedded within this study is an internal program evaluation consisting of multiple-choice questions and open-ended text-response questions. Specifically, upon program exit (data collection time two) parents will be asked to complete a five-minute survey relating to program usability and program satisfaction. Example questions asked include: 1) *How useful was the program for you, as a parent of a young child?* and 2) *What was most helpful about* MERTIL *for Parents?* This will be sent to parents via email with an attached secure link.

#### Nurses

After hearing from their parent participants, nurses will be asked to complete a brief mixed-method online survey, allowing insight into nurse perceptions of the program. Questions will include: 1) *Would you recommend MERTIL for Parents to future clients/patients?* 2) *Would you recommend colleagues encourage suitable clients/patients to participant in MERTIL for Parents?* 3) *From your perspective, does MERTIL for Parents fill a current preventative health gap?* 4) *Are there any existing gaps in the MERTIL for Parents program?* For each question response options will be ‘yes’ or ‘no’, and then participants will be asked to expand upon their answer with free text.

### Focus groups

To further examine the feasibility and efficacy of MERTIL *for Parents*, focus groups will be held upon program completion. Each group will consist of 4-6 parents or nurses. A semi-structured interview schedule will guide focus group discussions. All focus groups will be audio recorded to allow for later transcription and analysis.

#### Nurse

Example focus group questions will include: 1) *Could you tell us about your MERTIL for Parents experience?* 2) *What was most relevant to you?* 3) *Can you see yourself in the future encouraging suitable parents to participate in MERTIL for Parents as a first intervention step?* (3) *What are the long-term and short-term implications of programs such as MERTIL for Parents*?

#### Parent

Example focus group questions will include: 1) *Could you tell us what it was like to participate in MERTIL for Parents?* 2) *Since completing MERTIL for Parents, what, if anything, has changed in your thinking, or feeling, or in your behaviour as a parent?* 3) *In MERTIL for Parents, what was most relevant to you?* 4) *Would you recommend MERTIL for Parents to other parents?*

### Program data management

All MCH nurse and parent survey data will be disseminated through Moodle, an online learning management system stored on a secure server, with data transferred securely for analyses. All study data and participant information will be stored securely as per the criteria required by the La Trobe University Human Ethics Committee.

### Data analysis strategy

#### Quantitative

All statistical analyses will be conducted using the software IBM SSPS Statistics (43). Descriptive statistics will be generated to identify nurse and parent population characteristics and to evaluate parent and nurse program perceptions. To determine parenting change in dyad attachment, relational function, maternal mental health, from pre-program to three-months post-program, inferential statistics will be used (i.e., univariate, bivariate, and multivariate analyses) such as t-tests, chi-squared analyses, ANOVA, and regression analyses.

#### Qualitative

##### Focus groups

All parent and nurse focus group interview data will be audio-recorded, transcribed verbatim, and deidentified. Thematic and content analysis will be used to identify themes and sub-themes from MCH nurse and parent transcript responses (44).

##### Free-text surveys responses

For any nurse or parent free-text survey responses, thematic analysis will also be used to identify response themes and sub-themes.

### Data collection commencement

Pilot field studies, including recruitment and data collection, will commence in August 2022 with data collection concluding in November 2022.

## Discussion

### Limitations and challenges

While this study is innovative, we foresee several program content and delivery method limitations: Firstly, it is possible that the brief self-guided content of MERTIL *for Parents* may not provide parents with adequate content information and support (i.e., program dose too low). Second, as a frontline service, MCH nurses may lack the time in session to explain and recommend suitable parents to participate in MERTIL *for Parents*, due to competing demands and scheduling pressures. Third, as the study population pertains only to nurses and parents located in Victoria and New South Wales, Australia, generalisability concerns will be present for inter-state Australian and international populations. Fourth, as all MERTIL *for Parents* content is housed online, this may exclude those who lack the technology or skills to participate, especially as on-demand technical support will not be available for live participant trouble shooting. Further, because of this study’s non-randomised sampling strategy, cause-effect relationships cannot be drawn. That is, parents’ socioemotional outcome changes cannot be causally inferred by their MERTIL *for Parents* participation. Finally, as with all research, there is a risk that attrition rates may be high. All parent data will be self-report in nature, and interpretation of findings will therefore be subject to these limitations.

### Strengths and opportunities

There are multiple strengths to the present study: First, the mixed-methods design of this study allows for greater understanding of the program’s impact compared with either qualitative- or quantitative-only designs. Second, diversity of parent demographics and needs will be ensured through recruitment of participants from both the Universal and Enhanced MCH services in both metropolitan and regional locations. Third, socioemotional-specific content may fill an important gap in the available online parenting program market. Fourth, the program’s reach is expected to be wide due to the online delivery method. Finally, strong completion is expected due to 1) the program’s brief nature, 2) the engaging, non-judgemental, and supportive program content; and 3) the contribution of MCH nurses, infant mental health experts, and parents in co-designing the program.

## Conclusion

This study will examine implementation feasibility and efficacy of an evidence-based, online, universal, self-directed parenting program, MERTIL *for Parents*. Multi-informant data from parents and nurses will inform future iterations of MERTIL *for Parents* and universal online relational programs.

## Data Availability

No datasets were generated or analysed during the current study. All relevant data from this study will be made available upon study completion by contacting the first author, JO.

## Acknowledgements

We wish to thank the maternal child health nurses, early childhood mental health clinicians, and parents who co-designed MERTIL *for Parents*.

## Data availability statement

Not applicable as data collection has not commenced. Once all study data is collected, the data supporting the findings of this study will be available from the corresponding author, JO, upon reasonable request.

